# Metastatic risk stratification of leiomyosarcoma patients using transcription- and replication-associated chromosomal instability mechanisms

**DOI:** 10.1101/2021.01.29.21250218

**Authors:** Ataaillah Benhaddou, Laetitia Gaston, Gaëlle Pérot, Nelly Desplat, Laura Leroy, Sophie Le Guellec, Mohamed Ben Haddou, Philippe Rochaix, Thibaud Valentin, Gwenaël Ferron, Christine Chevreau, Binh Bui, Eberhard Stoeckle, Axel Le Cesne, Sophie Piperno-Neumann, Françoise Colin, Nelly Firmin, Gonzague De Pinieux, Jean-Michel Coindre, Jean-Yves Blay, Frédéric Chibon

## Abstract

Leiomyosarcoma (LMS) is an aggressive smooth muscle cancer with few therapeutic options. LMSs show a high level of genomic instability (GI) and the mechanisms underlying their oncogenic processes are poorly understood. While the level of GI influences treatment efficacy and resistance, an accurate measure of it is lacking. Current measures of GI are based on counts of specific structural variation (SV) and mutational signatures. Here, we present a holistic approach to measuring GI based on the quantification of the steady-state equilibrium between DNA damage and repair as assessed by the residual breakpoints (BP) remaining after repair, irrespective of SV type. We use the notion of Hscore, a BP “hotspotness” magnitude scale, to measure the propensity of genomic structural or functional DNA elements to break more than expected by chance. We then derived new measures of transcription- and replication-associated GI that we call iTRAC (Transcription-Associated Chromosomal instability index (iTRAC) and iRACIN (Replication-Associated Chromosomal INstability index). We show that iTRAC and iRACIN are predictive of metastatic relapse in LMS and that they may be combined to form a new classifier called MAGIC (Mixed transcription-and replication-Associated Genomic Instability Classifier). MAGIC outperforms the gold standards FNCLCC and CINSARC in stratifying metastatic risk in LMS. Furthermore, iTRAC stratifies chemotherapeutic response in LMS. We finally show that this approach is applicable to other cancers.

## Introduction

Leiomyosarcoma (LMS) with smooth muscle cell (SMC) differentiation is one of the most frequent soft tissue sarcomas (STS), a group of tumors that arise from connective tissue cells. However, LMS is rare, with about 600 cases in France every year (Blay et al. 2017). While LMS can occur at any anatomical site like other STS, the limbs, retroperitoneum, and uterus are the three main locations. The standard treatment for patients with localized LMS is wide surgical resection to obtain tumor-free margins. LMS is a highly aggressive STS subtype and as many as 50% of patients relapse. In this setting on an incurable disease, median survival is 12 months (Judson et al. 2014). Since neither targeted therapy (van der Graaf et al. 2012) nor immunotherapy (Ben-Ami et al. 2017) have demonstrated major therapeutic effects in LMS so far, anthracycline-based chemotherapy is the treatment of choice for metastatic LMS.

LMS develops due to frequent p53 and RB1 pathway alterations (Chibon et al. 2010; Derré et al. 2001) and a highly rearranged genome with a high number of chromosomal rearrangements. This leads to many copy number variations (CNV) and BP that are associated with poor outcome (Pérot et al. 2010). The stratification of LMS has long been based on histological measures like FNCLCC grading (Coindre et al. 2001) and is currently challenged by expression-based signatures (Chibon et al. 2010). Next-generation sequencing (NGS) has recently demonstrated the ability to identify clinically actionable genetic variants across many genes (Spencer et al. 2013). Current approaches in cancer genomics often use exome-seq to build a catalogue of mutations in multiple cancer types by sequencing hundreds of tumor samples in order to find diagnostic, prognostic, and therapeutic targets (Shabani Azim et al. 2018). While this approach remains relevant in most cancer types, it is rapidly becoming insufficient in highly rearranged cancer types like LMS, where recurrent driver gene mutations are very rare (Andersson et al. 2016) and it is more likely that their rearranged genome is actually the driving force of oncogenesis (Davoli et al. 2017). Here we used whole genome sequencing (WGS) of LMS tumor samples to identify SV across all tumor genomes and to infer the mechanisms of GI at the genome level.

GI is a hallmark of cancer (Negrini, Gorgoulis, et Halazonetis 2010) and may arise due to deleterious mutations in components of DNA repair pathways or to abnormally high levels of genotoxic stress from cellular processes such as transcription and replication that overwhelm high-fidelity DNA repair (Tubbs et Nussenzweig 2017). Replication stress is a threat to genome stability and has been implicated in tumorigenesis (Gaillard, García-Muse, et Aguilera 2015; Macheret et Halazonetis 2015; Técher et al. 2017). Notably, common fragile sites in cancer colocalize with chromosomal regions that are particularly prone to breakage following mild replication stress (Debatisse et al. 2012; Le Tallec et al. 2013; Blin et al. 2019). Transcription also creates conditions for mutations and recombination as well as DNA breaks, either by transcription-associated processes or by its ability to become a barrier to DNA replication (Aguilera 2002; Jinks-Robertson et Bhagwat 2014; Kim et Jinks-Robertson 2012; Gaillard, Herrera-Moyano, et Aguilera 2013; Marnef, Cohen, et Legube 2017). Indeed, co-transcriptional R-loops constitute a barrier for replication fork progression and lead to fork stalling and collapse. This is thought to be a major mechanism of GI that involves transcription-replication collisions (Helmrich, Ballarino, et Tora 2011; Wilson et al. 2015; Pentzold et al. 2018; Madireddy et al. 2016). Finally, obstacles on the template DNA, such as non-B DNA (NBD) structures, DNA repeats, DNA-bound non-histone proteins and transcription complexes, can impede replication fork progression (Gaillard et Aguilera 2016; Azvolinsky et al. 2009; French 1992; Deshpande et Newlon 1996; Gómez-González et al. 2011).

GI has long been thought to be predictive of poor prognosis, although an accurate method for measuring it remains to be discovered (Ahmad, Ahmed, et Venkitaraman 2018). Furthermore, the current lack of a measure that captures the dynamic nature of GI limits our ability to leverage it for prognostic and therapeutic purposes (Sansregret, Vanhaesebroeck, et Swanton 2018) in a clinical setting. We therefore sought to evaluate whether transcription complexes, as well as NBD and DNA repeats, play any role in LMS GI. To do so, we used WGS data to compare LMS structural variations (SV) and BP to known sites of transcription complexes, NBD and DNA repeats, and evaluated whether these BP are enriched more than expected. We present the Hscore, a new Hotspotness magnitude scale. The Hscore measures the propensity of a given DNA element to break more than expected with a random breakage model. By using this approach, we have developed a combination of both transcription-associated and replication-associated markers of GI and tested whether both measures are prognostic of metastatic risk in LMS. Furthermore, we assessed whether the transcription- and replication-associated markers are predictive of chemotherapeutic response in LMS.

## Results

Whole genome sequencing of 112 LMS (supplemental table 1, supplemental table 2) with our homemade SV detection pipeline (see material and methods) allowed us to identify 24870 BP forming 12435 SV. Each SV is formed by 2 BP. Of all BP, 67.4 % (16764 BP) are implicated in intra-chromosomal SV (BPSVintra) and 32.6 % (8106 BP) in inter-chromosomal SV (BPSVinter). While most BP occur inside regulatory DNA elements (13377/24870 = 53.79 %), some affect NBD (4902/24870 = 19.71%) and others (3574/24870 = 14.37% of BP) arise in DNA repeats. Taken together, a total of 66.4% (16510/24870) of LMS BP were identified in the DNA elements in this study.

The total BP count per LMS (TBPc) was highly skewed (ranging from 26 to 1200; mean = 222.1, median = 181; fig 1A) and did not fit a normal distribution (Shapiro-Wilk test P = 1.4 × 10^−12^; fig 1A), but TBPc follows a log-normal distribution (Shapiro-Wilk test P = 0.81; fig 1B). This is not surprising since log-normal distributions are common in nature and reflect forces acting independently and whose interactions result in multiplicative effects (Limpert, Stahel, et Abbt 2001). In cancer, these forces may include genetic, environmental, physiological, immune, sub-cellular and supra-cellular constraints.

**Figure 1.**
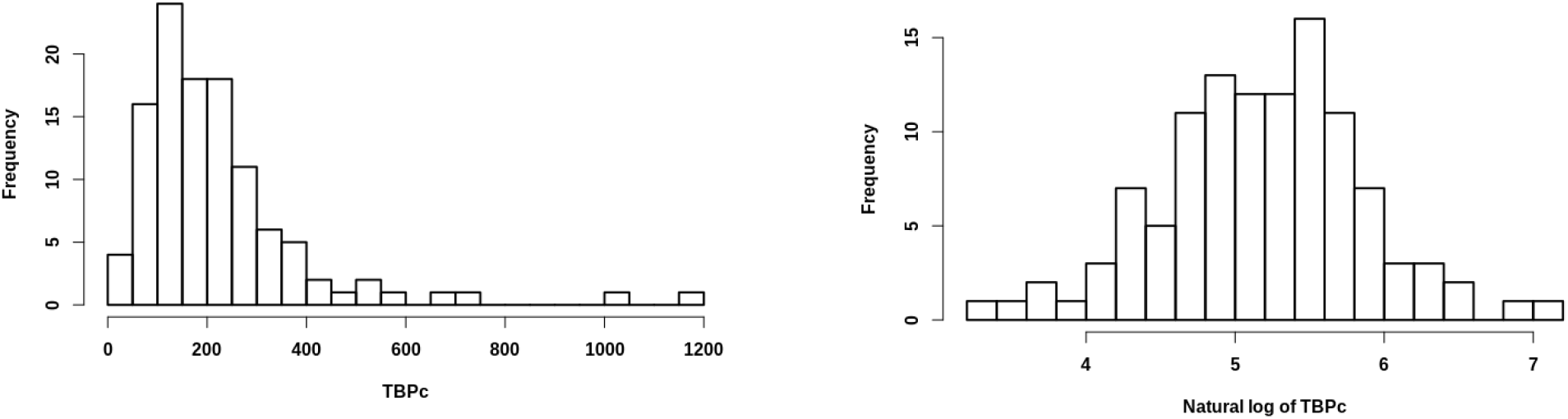
LMS BP are log-normal distributed. Left, histogram of TBPc. Right, histogram of natural log of TBPc.

Since the LMS genome is highly rearranged, the question arises whether DNA breakages occur randomly or are due to mechanisms associated with these constraints and are thus specific to certain regions of the genome. To address this question, we tested the “hotspotness” of three different types of genomic DNA structure: regulatory DNA elements, and NBD and DNA repeats. We propose that the Hscore may be used to assess the propensity of DNA elements to break more than expected by chance under the random breakage model (RBM) (fig 2A). The Hscore for a given type of DNA element is defined as the -log10 probability of having more BP than those expected in those elements, given the total number of BP, the cumulative size of the DNA elements and the readable genome size (materials and methods). The Hscore is very intuitive: the higher it is, the more significantly a given DNA element is more broken than expected. To determine whether a given genomic feature is a hotspot, we computed the Hscore in sliding windows upstream and downstream and made profile plots (fig 2A; see material and methods). Two significant broken structures emerged: hotspots and hot regions. We define the characteristics of hotspots as follows: 1) DNA elements are significantly more broken than expected by chance (Hscore ≥3) while the immediate surrounding regions are not (Hscore < 3); 2) if the DNA elements and the immediate surrounding regions have a Hscore ≥3, a DNA element is considered as a hotspot if its Hscore is at least 1.5 times the Hscore of both surrounding regions. A hot region is inferred if an Hscore ≥3 is observed for both the DNA element and its surrounding regions, provided that the Hscore of the DNA element is less than 1.5 times the Hscore of the surrounding regions.

**Figure 2.**
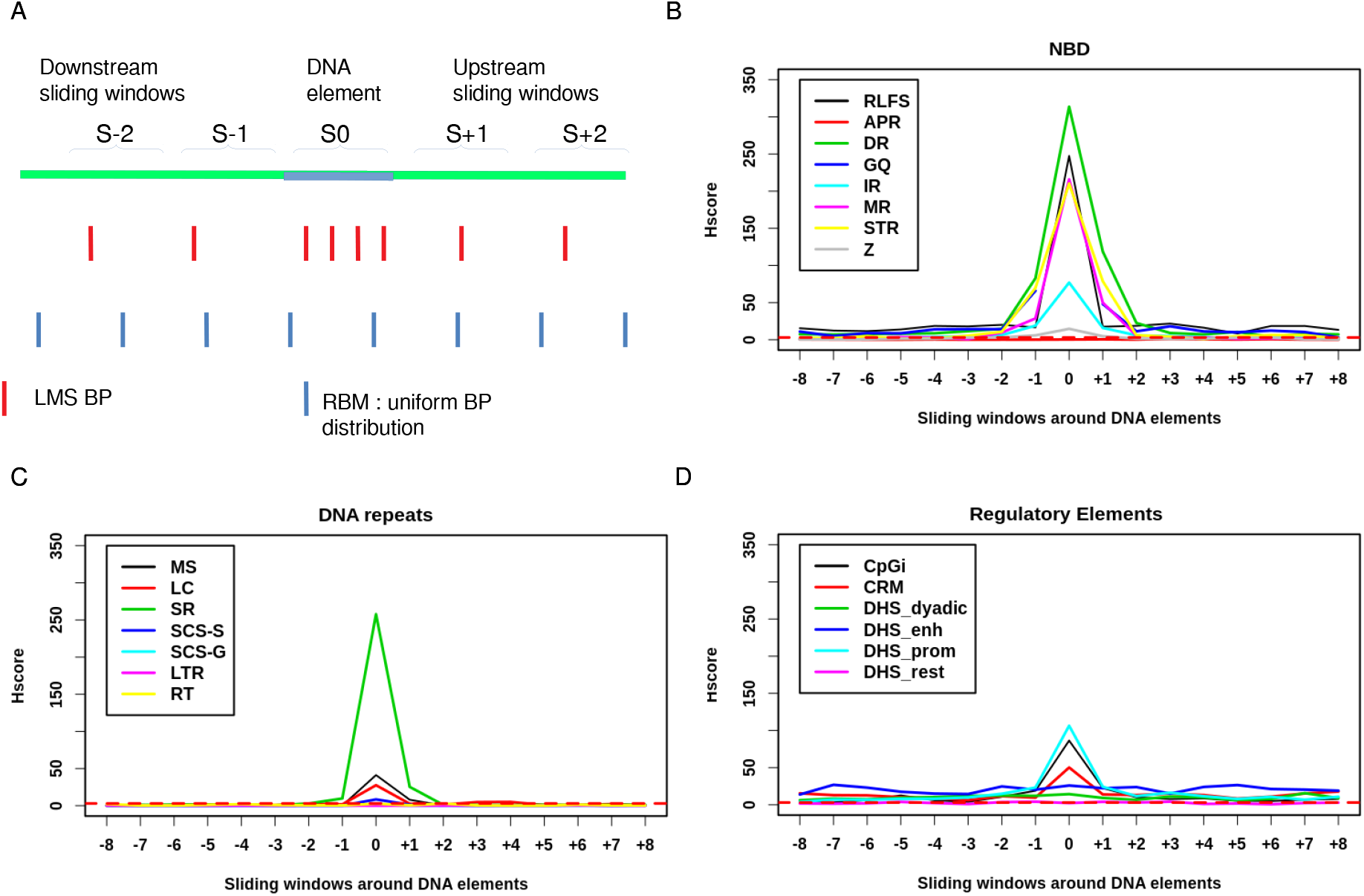
Regulatory elements, non-B DNA (NBD) and DNA repeats are hotspots. A. Random Breakage Model : hotspots are defined as DNA elements containing more BP than expected under RBM and more than surrounding regions. Hscore profile for DNA elements and sliding windows upstream and downstream for B. Non-B DNA and C. DNA repeats D. regulatory elements. Horizontal red dashed line corresponds to Hscore threshold of 3 for hotspotness.

### NBD and DNA repeats are hotspots for DNA breakage in LMS

NBD are DNA elements that adopt non-canonical DNA structures (Gaillard et Aguilera 2016). Sequences prone to form NBD are widespread in the human genome and are associated with GI (Wang et Vasquez 2014). The formation of NBD requires unwinding of the DNA sequence, as occurs during replication and transcription (Gaillard et Aguilera 2016). NBD comprise A-Phased Repeats (APR), Direct Repeats (DR), G-Quadruplex (GQ), Inverted Repeats (IR), Mirror Repeats (MR), Short Tandem Repeats (STR), Z DNA (Z) and RLoops Forming Sequences (RLFS). We found that all NBD excpet APR are hotspots (fig 2B), especially GQ which are so highly broken so that the Hscore tends to infinity (P = 0).

The DNA repeats investigated in this study comprise high-copy repeats: MicroSatellite (MS), Low Complexity (LC), simple repeats (SR); and low copy repeats like Self Chain Segments (SCS), which we split into self-aligned (SCS-S) and gapped (SCS-G), Long Terminal Repeats (LTR) and Retro Transposons (RT). While SR, MS, LC are hotspots, the viral origin repeats LTR and RT are not (fig 2C). Concordantly, no viral insertion was observed in LMS genomes when using HGT tools (Nguyen et al. 2015 and data not shown). Furthermore, while SCS-S were hotspots, SCS-G were not.

### Genes promoters are hotspots; gene enhancers are hot regions

The promoters of transcriptionally active genes have repeatedly been shown to recurrently harbor double-strand breaks (DSB) (Marnef, Cohen, et Legube 2017). Furthermore, DNase Hyper sensitive (DHS) DNA elements as well as active chromatin marks have been shown to colocalize with DSB (Mourad et al. 2018). The regulatory DNA elements used in this study comprise CpG islands (CpGi), Cis regulatory Modules (CRM), and DHS of promoter type (DHS_prom), of enhancer type (DHS_enh), of dyadic type (both enhancer and promoter signatures) (DHS_dyadic), and of other types (DHS_rest) (Roadmap Epigenomics Consortium et al. 2015). We found that promoter-associated DNA elements (*i.e*. DHS_prom and CpGi) are hotspots for DNA breakage (fig 2D) while DNA elements associated with enhancer activity (DHS_Enh) are hot regions (fig 2D). Furthermore, promoter-associated regulatory elements (CpGi, DHS_prom) break more significantly than Enhancers (DHS_enh), which break more significantly than Dyadic regulatory elements (DHS_dyadic) and the rest of DHS (DHS_rest) (fig 2D). Interestingly, CRM, which comprises overlapping regulatory regions (promoters, enhancers, dyadic) (Chèneby et al. 2018), is a hotspot and has an intermediate Hscore. This further underlines the robustness of our BP hotspotness magnitude scale. Finally, DHS which are not labeled to have any gene regulatory function (DNS_rest), are not significantly broken more than random (fig 2D). Taken together, these results show that DNA breakages are more enriched in promoters than in enhancer and dyadic regions. Furthermore, BP are more significantly enriched in open chromatin structures (DHS) with attributable gene regulatory functions like enhancers and promoters than DHS, with no attributable gene regulatory function, like DHS_res. Therefore, because DHS has an open DNA structure both inside and outside genes, we hypothesize that they do not break merely due to the openness of their chromatin but rather due to their genic context. We thus refined our analysis by splitting each DNA element into two categories: located inside or outside genes.

### Regulatory elements are almost exclusively “hot” inside genes and not outside them

To address the impact of the functional genomic context of a DNA element on the BP frequency, we split them into those located inside genes from the transcription start site (TSS) to the transcription end site (TES) (including DNA elements with at least 1 bp overlapping with genes) and those located outside genes, and then computed the Hscore for each group in sliding windows. We found that all regulatory elements (fig 3) were more significantly broken when they were located inside genes than outside, independently of their structure (sequence type like CpGi vs DNA-protein complexes) or function (promoter type vs enhancer type). CRM were always broken more frequently than by chance when located inside genes, with an Hscore of 67.82 inside genes and less than 3 outside (fig 3A, supplemental table 3). Although CpGi located outside genes were slightly significantly broken (Hscore=6.41), this significance was much higher when they were located inside the genes (Hscore=83,30) (fig 3B), with a ratio of Hscores inside/outside genes (RHscore i/o) of 12.99 (supplemental table 3). Similarly, all types of DHS regulatory elements were broken more significantly inside genes than outside (fig 3 C, D, E and F). These results suggest that the transcriptional and/or epigenetic context of genes strongly modulates the ability of DNA regulatory elements to harbor DNA BP. Since regulatory elements have an open DNA structure both inside and outside genes when they are active, we rule out the possibility that their propensity to harbor DNA BP is merely due to the openness of their chromatin.

**Figure 3.**
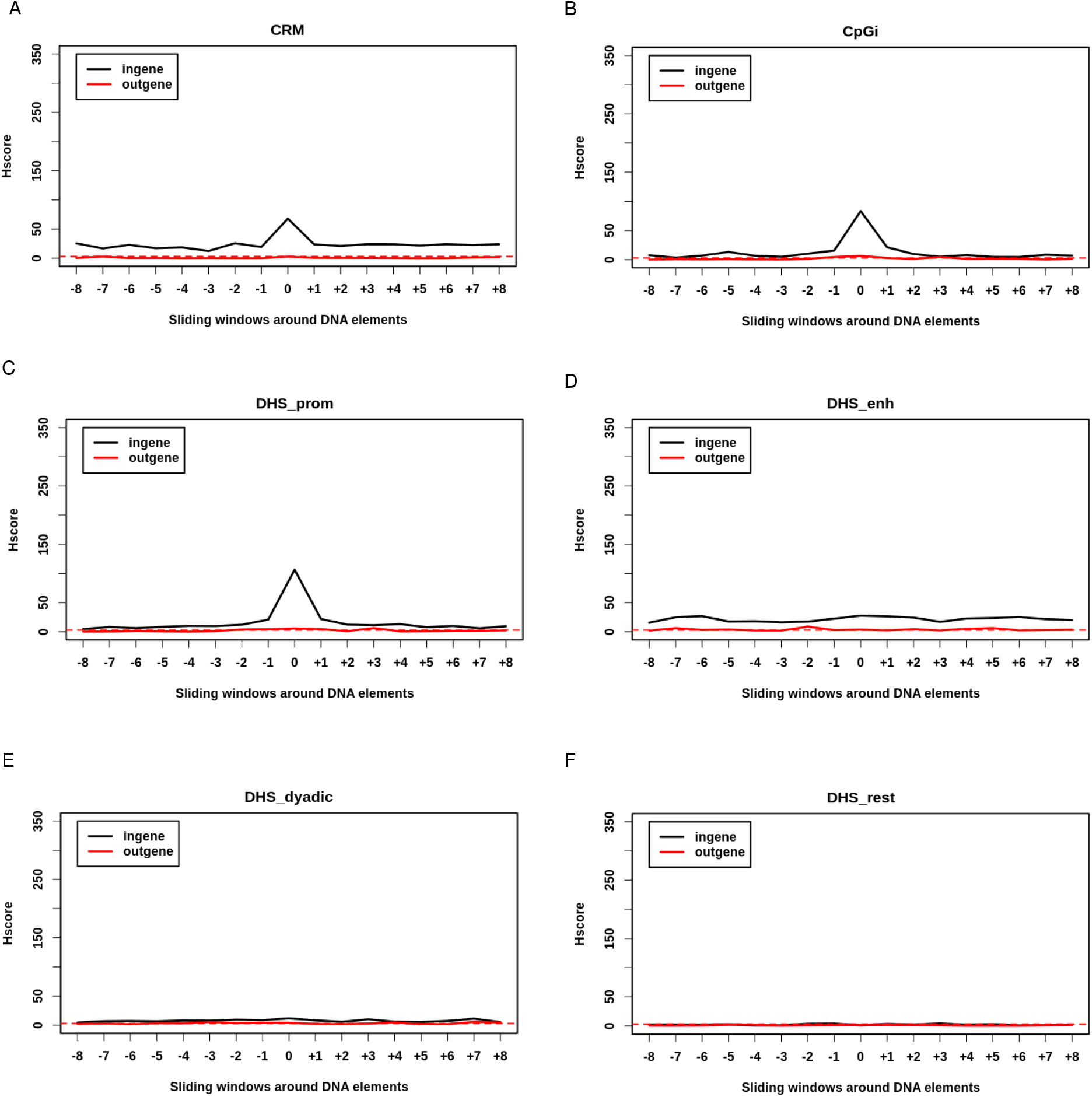
Regulatory elements are exclusively hot inside genes : Hscore for regulatory elements and sliding windows either inside (black) or outside genes (red). A, Cis-Regulatory Modules (CRM). B,CpG islands (CpGi). C, DNase Hyper Sensitive sites (DHS) of type promoter. D, DHS of type enhancer. E, DHS of type dyadic. F, DHS of other types. Horizontal red dashed line corresponds to Hscore threshold of 3 for hotspotness.

### NBD and DNA repeats are hotspots both inside and outside genes

We also split NBD and DNA repeats into those located inside genes and those located outside them (fig 4). MR, IR, DR, and STR were significantly broken to a similar extent inside and outside genes (Ratio Hscore i/o 1.02, 1.12, 1.02, 1.13 respectively) (fig 4A, B, C and D). While RLFS were highly significantly enriched in BP outside genes (Hscore=66.24), they were even more significantly broken inside them (fig 4E) (Hscore=185.41, RHscore i/o=2.8) (supplemental table 3). GQ were also highly significantly enriched in BP outside genes (Hscore=180.53) and tended to be more broken inside them (Hscore=230.17, RHscore i/o=1.27) (fig 4F). Z DNA tended to be more frequently broken outside genes than inside them (RHscore i/o = 0.73) (fig. 4G). Hscores of DNA repeats located inside and outside genes were almost identical except for SCS-S, which were significantly more broken than random inside genes (Hscore=8.94) but not outside them (Hscore=1.18<3), while SCS-G were indistinguishable from random (fig. 5). Together these results suggest that, except for SCS-S, GQ and RLFS, NBD and DNA repeats are insensitive to gene context and may cause GI by a mechanism largely independent from transcription and which is probably replication-dependent. Interestingly, GQ and RLFS share the properties of the three DNA element types: they are sensitive to their genic context as regulatory elements and are breakable when located outside genes, as are DNA repeats and NBD.

**Figure 4.**
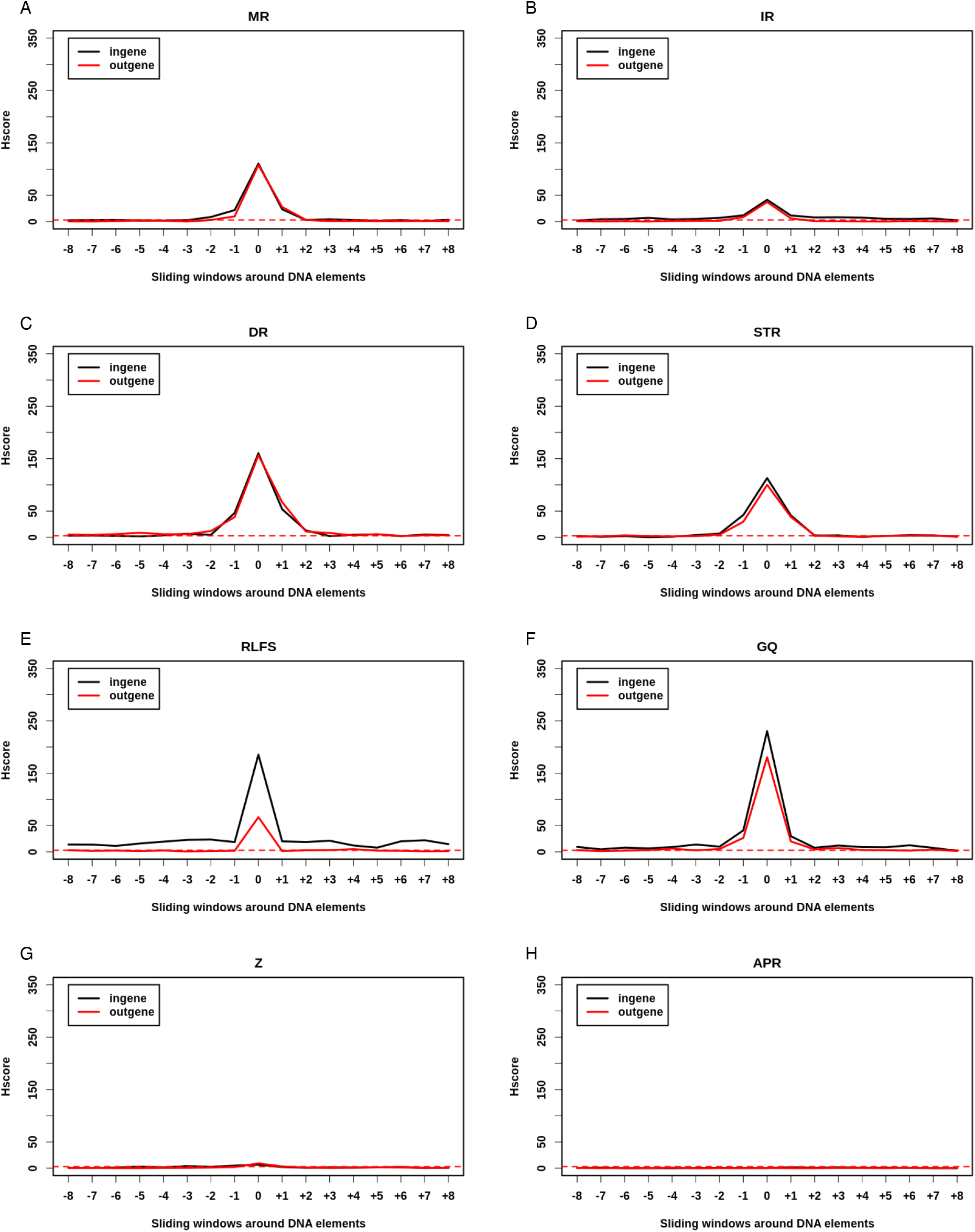
NBD are hotspots both inside and outside genes : Hscore for NBD and sliding windows either inside (black) or outside (red) genes. A, Mirror Repeats. B, Inverted repeats. C, Direct Repeats. D. Short Tandem Repeats. E. R-loops forming sequences. F. G-quadruplex. G, Z DNA. H. A-phased Repeats. Horizontal red dashed line corresponds to Hscore threshold of 3 for hotspotness.

**Figure 5.**
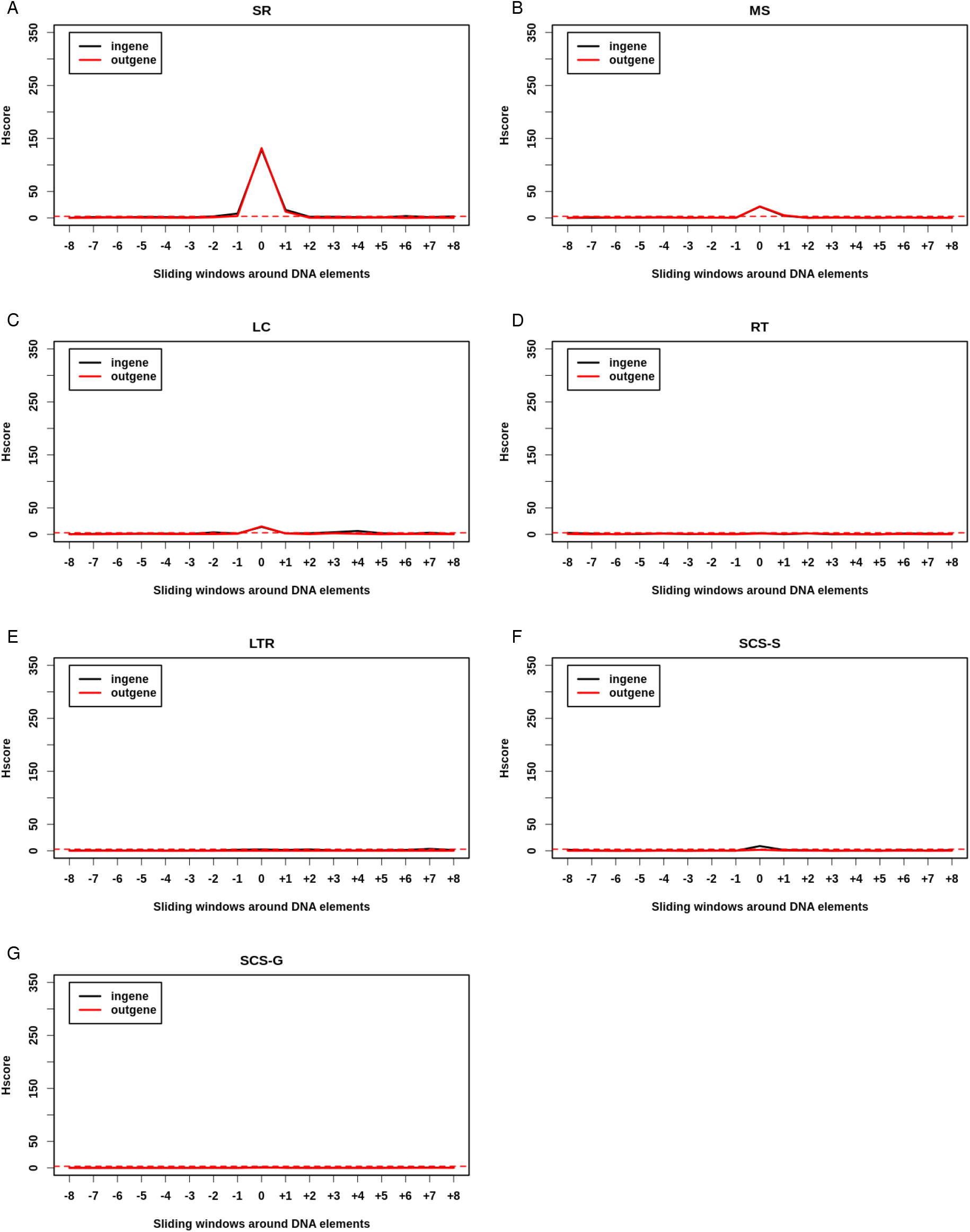
DNA repeats hotspotness relative to genes is dependent upon their type. High-copy DNA repeats : A-C. A, Simple Repeats. B, MiscoSatellites. C, Low Copy repeats. Viral orgin DNA repeats : D, Retro-Transposons. E, Long Terminal Repeats. Low copy repeats : Selfchains F-G .F, Selfchains self aligned. G, Self chain regions. Horizontal red dashed line corresponds to Hscore threshold of 3 for hotspotness.

### Not all LMS present BP distributed as hotspots

The abovementioned results obtained on the whole cohort were global so we still did not know what happens in each patient. To evaluate the DNA breakage mechanisms present in each patient, we computed the Hscore for regulatory elements, NBD and DNA repeats in each LMS tumor sample and made a hierarchical clustering of LMS patients based on these Hscores (fig 6, see material and methods). We found that not all LMS patients had BP distributed as hotspots and that there was a gradient of hotspotness. Furthermore, each LMS sample had its specific profile: while some patients had no detectable hotspots, others had BP hotspots mainly in the regulatory regions, and others still had them mainly in NBD and DNA repeats. There were even some patients who had them in both (fig 6). Interestingly, metastatic and non-metastatic patients were not evenly distributed over the gradient of hotspotness, the hotspot side of the heatmap having fewer metastatic events than the opposite side (fig 6). We therefore address the question of the relation between BP hotspotness and patient prognosis with different approaches in the next sections.

### The LMS cohort can be stratified into clinically relevant groups

Because a) regulatory elements, NBD and DNA repeats have been thoroughly documented to impede transcription and replication in both a transcription-associated (Gaillard et Aguilera 2016) and replication-associated (Gaillard, García-Muse, et Aguilera 2015) manner and to cause GI, and b) GI is predictive of poor prognosis (Ahmad, Ahmed, et Venkitaraman 2018), we hypothesized that there is a link between transcription- and replication-associated DNA-breakage mechanisms and metastatic clinical outcome in LMS. To test this hypothesis, we sought to quantify the overall transcription-associated and replication-associated GI. We used our BP hotspotness magnitude scale and derived genomic indexes for both transcription-dependent and transcription-independent DNA-breakage and genome instability. As DR, STR, MR, IR, Z DNA, SR, MS and LC are BP-enriched irrespective of their position inside or outside genes, we considered these elements as transcription-independent and thus as Replication-Associated Chromosomal INstability elements (RACINe). Conversely, we considered RLFS, GQ, CpGi, CRM, SCS-S and DHS as Transcription-Associated Chromosomal instability elements (TRACe), because they are more frequently broken than chance inside genes and not outside them. By consolidating TRACe and RACINe into one functional group and computing Hscores (see material and methods), we derived a TRAC index (iTRAC) and a RACIN index (iRACIN), respectively. iTRAC and iRACIN allow the quantification of the overall contribution of RACINe and TRACe to DNA breakage and therefore to GI in each LMS patient. To address the relationship between iTRAC, iRACIN and metastatic clinical outcome, we developed a method called Iterative multi-threshold PARTioning (iPART) (see material and methods).

First, we used iPART to establish a threshold for iTRAC and iRACIN that splits the LMS cohort into two groups with a maximum difference in MFS (Metastasis-Free Survival). Figure 7A and 7B left panels show that iTRAC has several thresholds giving p-values less than 0.05 (red horizontal dashed line), while iRACIN has only a few of them. We also noted the presence of several local minima that triggered, suggesting that there could be more than one threshold to split the LMS cohort into groups of different MFS. To test this idea, we applied iPART using all combinations of two thresholds from all local minima below 0.10 for iTRAC and 0.3 for iRACIN (blue horizontal dashed line) (materials and methods). Accordingly, both iTRAC and iRACIN significantly stratified the LMS cohort (P = 3.08 × 10^−5^ and P = 4.13 × 10^−5^, respectively) into three groups with distinct outcomes (fig 7A and 7B right panels). Strikingly, it was the groups of LMS with medium iTRAC or medium iRACIN which had the most unfavorable outcome (fig 7A and 7B right panels).

**Figure 7.**
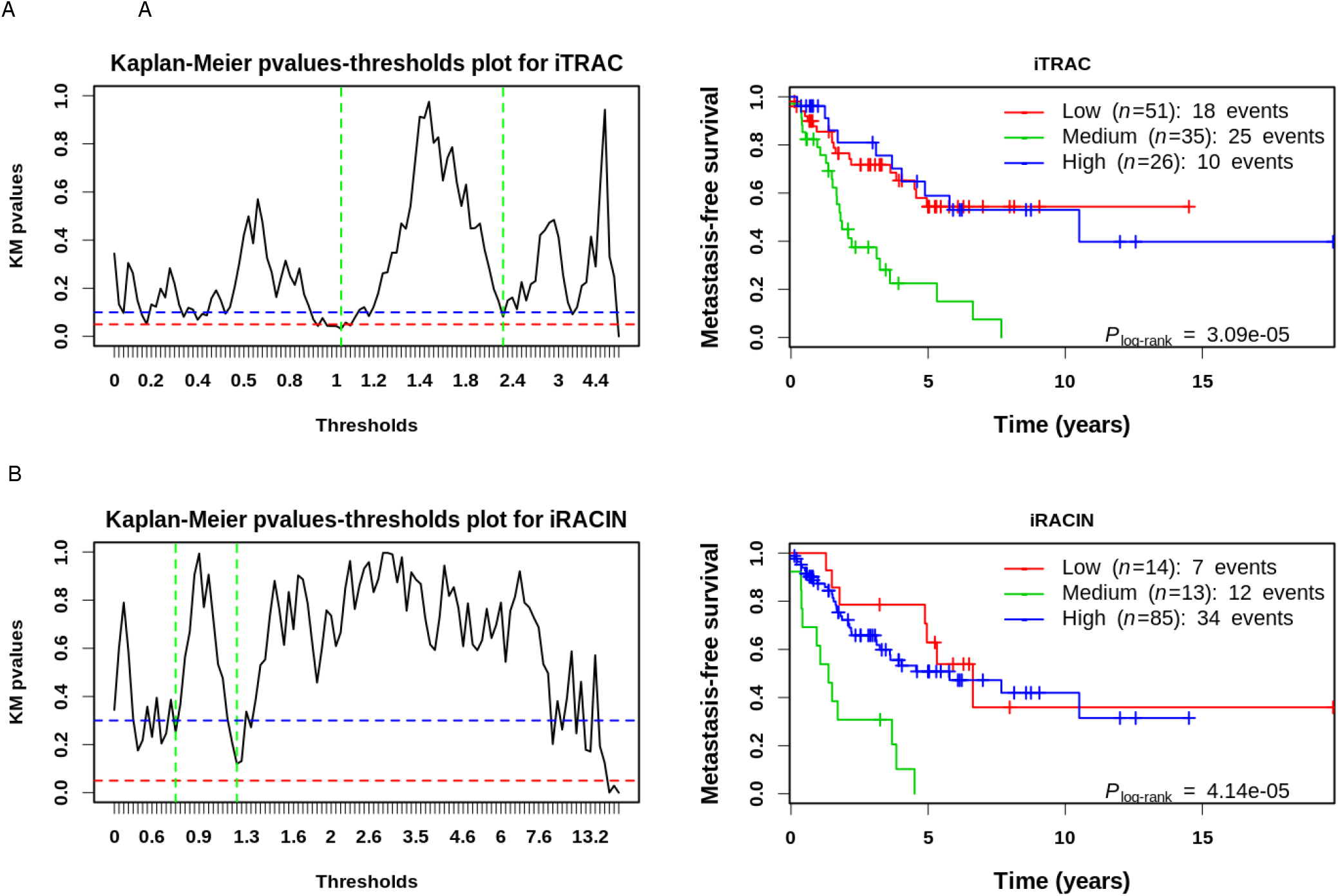
Stratification of the LMS cohort into Low, Medium and High levels of TRAC and RACIN using iPART. A iTRAC. Left, Kaplan-Meier pvalues in function of iTRAC single threshold dividing the LMS cohort into Low and High groups (black solid line). Red dashed horizontal line corresponds to 0.05 arbitrary and commonly accepted significance pvalue threshold. Blue dashed horizontal line corresponds to the pvalue threshold we considered for pvalues that will be included in combinatorial double threshold division of the LMS cohort into Low, Medium and High groups (material and methods for more details). Vertical green dashed lines correspond to the best combination of thresholds (tl=0.99, th=2.29) spliting the LMS cohort into Low, High, Medium groups. Right, Kaplan-Meier plot for iTRAC stratified LMS cohort into Low, Medium and High based on the best combination of thresholds tl=0.99 and th=2.29. B. iRACIN. Both plots correspond to the same procedure as A. tl and th for iRACIN are 0.74 and 1.30 respectively. tl, Treshold Low. th,Threshold High.

We also applied iPART on TBPc, the number of BP implicated in intra-chromosomal SV (nBPSVintra), the number of BP implicated in inter-chromosomal SV (nBPSVinter). Although TBPc, nBPSVintra and nBPSVinter were slightly significant (supplemental fig 1), they were not comparable to the level of significance we obtained by iTRAC and iRACIN. Thus, iTRAC and iRACIN have far more added value in the stratification of metastatic risk in LMS than mere BP counts, further stressing the relevance our approach.

### Mixed transcription and replication-Associated Genomic Instability Classifier (MAGIC)

To translate these results into a clinically relevant stratification tool, we sought to integrate both iTRAC and iRACIN into one classifier called the Mixed transcription- and replication-Associated Genomic Instability Classifier (MAGIC). Given that both indexes have high and comparable statistical significance in stratifying LMS, we considered at high risk (MAGIC High-risk) any patient classified as medium risk by either iTRAC or iRACIN, and the rest of patients as low risk (MAGIC Low-risk). MAGIC achieved a very high level of significance in stratifying LMS samples (P = 8.75 × 10^−8^; fig 8A), with a median MFS for the MAGIC High-risk group of 1.8 (CI = [1.52, 3.61]) which was 5 times lower than for MAGIC Low-risk group (10.5, CI= [5.78, NA]).

**Figure 8.**
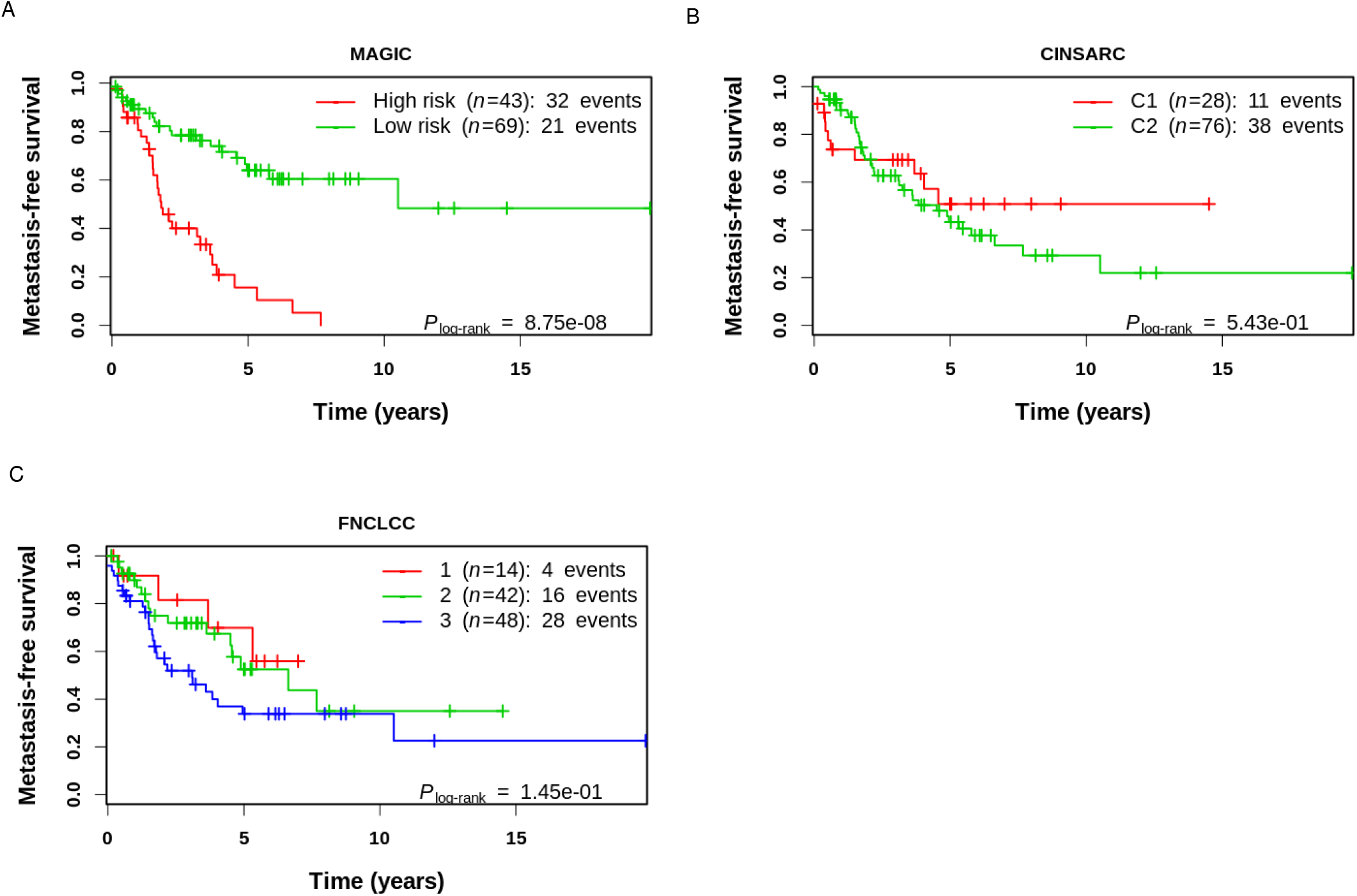
MAGIC outperforms CINSARC and FNCLCC in the stratification of metastatic risk in LMS cohort. Metastasis-free survival curves of LMS cohort stratified by A. MAGIC, B. CINSARC, C. FNCLCC

### MAGIC outperforms histologic FNCLCC and molecular CINSARC gradings

The histological FNCLCC grading system predicting patient evolution is the current standard in sarcomas (Coindre et al. 2001; Guillou et al. 1997). CINSARC, which is the best molecular signature of sarcomas, is challenging the histological gold standard and is currently under clinical investigation for stratification (Chibon et al. 2010). Both approaches individually did not significantly split the LMS cohort into groups with different metastatic evolution (fig 8). This finding precludes the introduction of either of these grading systems in multivariate analysis, so we conclude that MAGIC strongly outperforms both the FNCLCC grading system and CINSARC in LMS metastasis risk stratification.

### iPART stratifies a Pan-Cancer cohort of twelve cancer types significantly into clinically relevant groups

The intermediary level of GI resulting in poor clinical outcome compared to low and high levels was also reported in a Pan-Cancer study of 12 cancer types (TCGA cohort) (Andor et al. 2016). The authors used CNV abundance as a measure of GI and found that CNV affecting between 25 % and 75 % of a tumor’s meta-genome was predictive of poor survival. We thus hypothesized that what we observed in LMS might be a general mechanism associated with tumor aggressiveness. To tackle this question in a reasonable time scale with our computational resources, we used the iPART algorithm directly on CNV abundance from “Andor *et al* 2016” as a proxy for iTRAC/iRACIN. Those authors used the arbitrary and commonly used method of splitting the data into quartiles based on thresholds of 25 %, 50 %,and 75 % to segment the cohort into four groups based on CNV abundance in the tumor meta-genome. Using iPART, we split their cohort into 2, 3, 4, 5, 6, 7 and 8 groups and evaluated the influence of each split on the risk of mortality using the Log-rank test and hazard ratio (Fig 9, supplemental Fig 2). The best data segmentation corresponded to 5 groups split with the log-rank test P = 6.9 × 10^−10^ and maximal HR = 4.7(p=3.42×10^−9^) (supplemental Fig 2). On the other hand, the data segmentation applied by Andor *et al*. had a log-rank test P = 5 × 10^−6^ (Fig.5b from Andor *et al*) and a maximum HR no exceeding 1.9 with P <0.005 (Fig.5c from Andor *et al*.). Thus, as Andor *et al*. showed previously, it is the intermediate group (G.3) which is associated with the worst overall survival and presents the highest HR (fig. 9; HR=4.75, p=3.42e-9). Since these results strengthened the robustness of our approach, we compared the prognostic value of MAGIC to that of established sarcoma prognostic tools.

**Figure 9.**
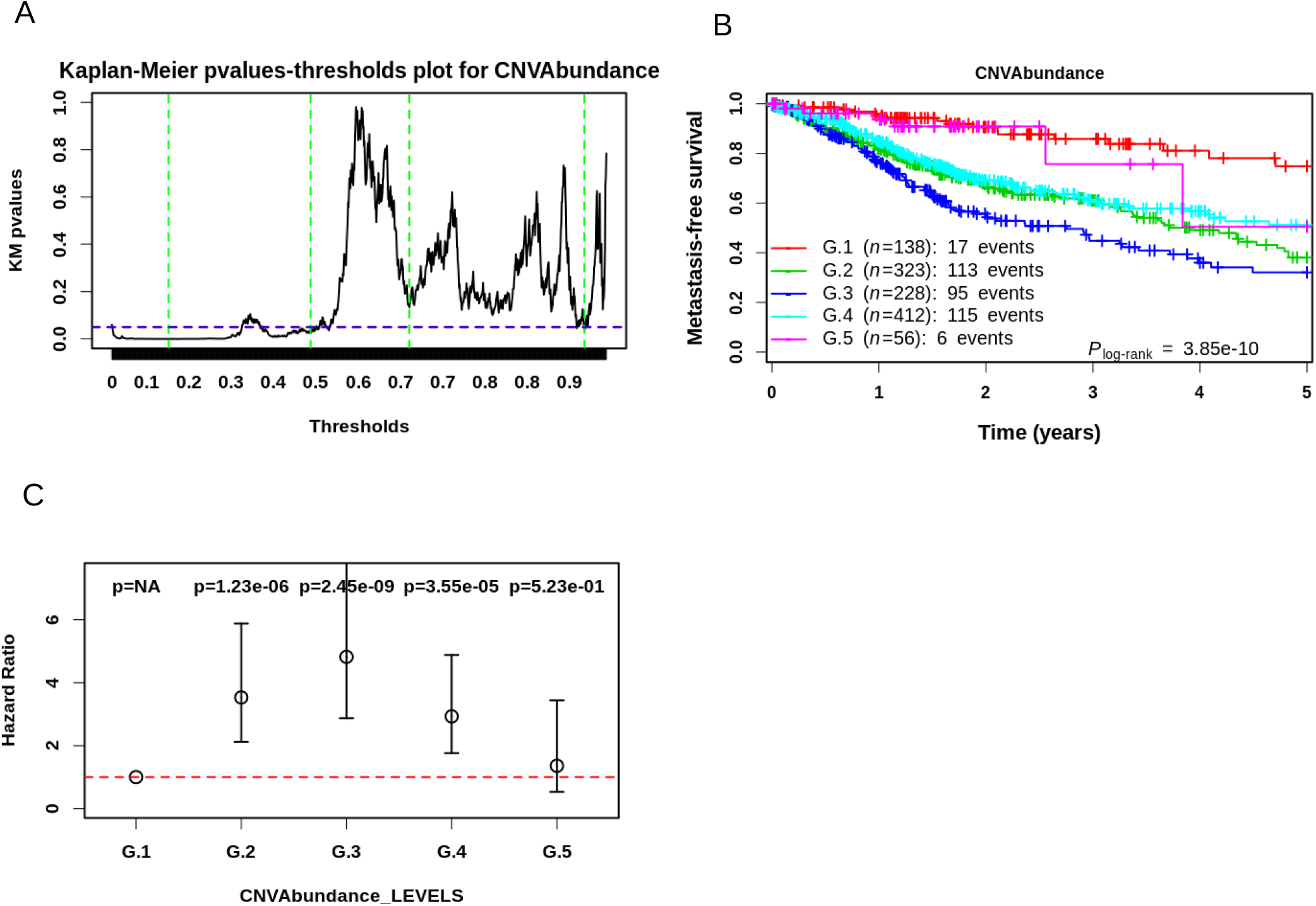
Application of iPART algorithm to the TCGA pan-cancer cohort of 12 cancer of Andor *et al*. 2016. A.Kaplan-Meier pvalues in function of CNVAbundance single threshold dividing the TCGA Pan-cancer cohort into Low and High groups (black solid line). Green vertical lines correspond to the four thresholds giving the best data segmentation (see text and methods for details) into 5 groups, which correspond from left to right to: 0.14, 0.45, 0.67, 0.86. B.Kaplan-Meier plot showing the stratification of the TCGA cohort into 5 clinically relevant groups. C.Hazard Ratio (HR) with 95 % confidence interval of risk of mortality of each group relative the reference group (G.1). Pvalues of each HR are shown above each condition. Horizontal dashed line corresponds to a Hazard ratio of 1.

### iTRAC stratifies chemotherapeutic response in LMS cohort

Chemotherapy remains controversial in LMS since no clinical trial has ever demonstrated its benefit. The criticism is that candidates, i.e. patients with a poor prognosis and responding to chemotherapy, are still not efficiently selected. We therefore sought to address this question in the 112 LMS in our cohort: 18 underwent chemotherapy (14 adjuvant, 1 neoadjuvant and 3 palliative), 18 patients were not annotated (NA) and 76 patients were not treated with chemotherapy. Because the level of GI influences the efficacy of several cancer treatments (Bakhoum et al. 2015; Janssen, Kops, et Medema 2009; Zasadil et al. 2014), we sought to quantify the contribution of TRAC and RACIN to the chemotherapeutic response in LMS. We first split the MAGIC risk groups according to their chemotherapeutic treatment status, *i.e*. Yes (18 pts) or No (76 pts). We found that MAGIC did not significantly stratify the chemotherapeutic response in LMS (fig 10A) as these patients, although similar in their metastatic risk, have rather different underlying oncogenic mechanisms. We then split each of the Low, Medium, and High groups of iTRAC, iRACIN according to chemotherapeutic treatment status. Interestingly, while patients in the Low risk iTRAC group receiving chemotherapy had a poorer prognosis (HR=4.47,CI=[1.65, 12.08],p=0.0032), no therapeutic benefit was found in the Medium and High risk iTRAC groups (fig 10A). In the High risk iTRAC group, there was a tendency for chemotherapy to be beneficial, but there was not enough statistical power (only three events in the chemotherapeutic arm) to test this hypothesis. Further patient inclusion would be needed to test the relevance of the prediction of the response to chemotherapy. Conversely, none of the iRACIN groups was relevant for stratifying chemotherapeutic response, again probably due to the small sample of treated patients (fig 10B).

**Figure 10.**
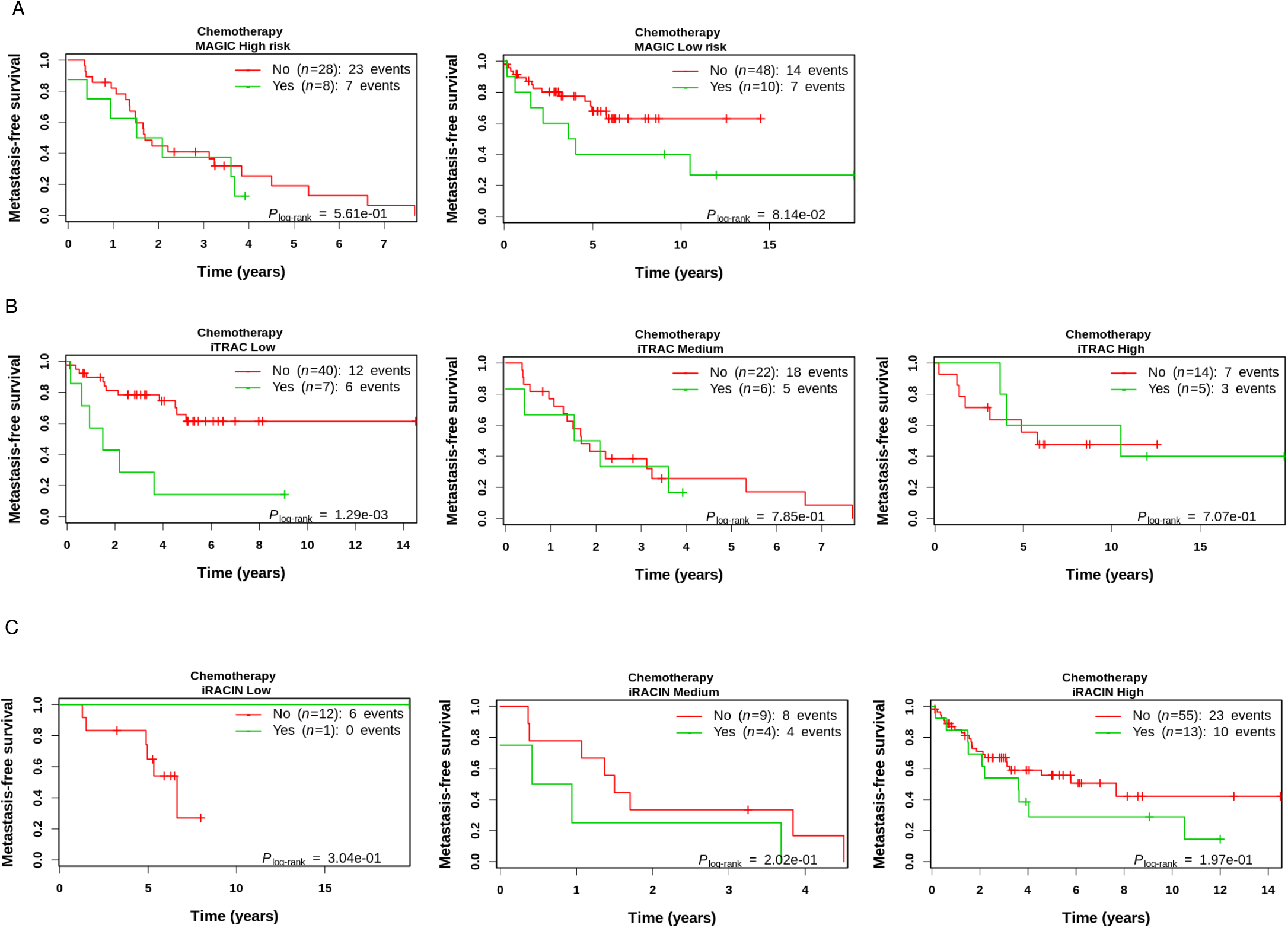
iTRAC stratifies chemotherapeutic response in LMS but not iRACIN. MFS curves in LMS groups of A. MAGIC, B. iTRAC and C. iRACIN stratified by chemotherapeutic treatment.

## Discussion

This study describes new tools, measures and insights that address the question of the clinical outcome of LMS and its relationship with genomic rearrangement. By deciphering the mechanisms of GI, we have produced the iTRAC and iRACIN indexes which account for transcription- and replication-related GI. We also generated the MAGIC classifier and demonstrated its prognostic value in outperforming both the current gold standard histologic and molecular challenger grading systems. Moreover, our indexes are potentially applicable to the Pan-Cancer cohort, as shown by the significant prognosis we established in twelve different cancer types using iPART and CNV abundance as a proxy for iTRAC/iRACIN.

### Application of Hscore on TRACe and RACINe as a holistic approach to measuring GI

Recent studies have used different GI scores and indexes to predict clinical outcome and to define homologous recombination (HR)-deficient samples (Zhang, Yuan, et Hao 2014; Birkbak et al. 2012; Abkevich et al. 2012; Popova et al. 2012; Stefansson et al. 2009; Baumbusch et al. 2013; Mirza et al. 2016). However, most of these studies were merely based on the per patient counts of *BRCA1/2* mutations, genome SNPs, loss of heterozygosity (LOH), or specific structural variations (CNV, Telomeric Allelic Imbalance, etc). These studies assume that an *a priori* selection of chosen genomic alteration types can recapitulate the dynamics of GI in a tumor that is complex, heterogeneous and subject to selection pressure from the tumor micro-environment. Here we present a new measure of GI that is based on the demonstration that GI is not a random process and is mainly due to the disruption of the steady-state equilibrium between continuous DNA damage and the matched level of high-fidelity repair maintaining genome integrity (Tubbs et Nussenzweig 2017). We introduce the notion of the Hscore, which is a BP hotspotness magnitude scale that measures the propensity of a given functional and/or structural genomic DNA element to harbor BP more than expected by chance. We also applied the Hscore on TRACe and RACINe as a holistic approach to measure GI based on measuring the steady-state equilibrium between DNA damage and repair. The method quantifies the residual DNA BP remaining after unsuccessful repair, irrespective of SV type, and quantifies the relative contribution of two main contributors to GI: TRAC and RACIN. The Hscore is measured on all tumor BPs without any biological-rational selection bias toward SV types. Values are comparable between different DNA elements in a patient and between patients. The higher the Hscore, the more unlikely it is that the observed BPs are due to random events. Therefore, the likelihood is greater that they are due to the structural and/or functional properties of those DNA elements.

### MAGIC: towards a new standard for LMS grading ?

LMS prognostication is still challenging and mandatory to evaluate which therapeutic strategy can benefit which patient. Since some patients have a poor prognosis associated with a transcription stress and others with a replication stress, we produced a simple patient-stratification tool called MAGIC. While neither histologic FNCLCC nor molecular CINSARC grading are suitable predictive methods in an LMS cohort, MAGIC achieved a high level of significance, with the high-risk group having a median MFS=1.8 years, i.e. five-fold lower than the low-risk group: 10.5 years. MAGIC could be used to spot patients with a high risk of metastasis. Prospective validation of this hypothesis is now mandatory.

### Prognostic relevance of iTRAC and iRACIN for metastatic risk stratification of other cancers

Unexpectedly, the risk of metastasis was found to follow a lambda (Λ) shape with the increase in iTRAC and iRACIN: both low and high GI levels correspond to a lower metastatic risk, while a medium level indicates a higher metastatic risk. Interestingly, similar results were previously reported in a Pan-Cancer analysis of 12 cancer types (Andor et al. 2016) concerning the abundance of copy number variations (CNV). Although the results were less statistically significant, the study highlighted the added value of quantifying transcription- and replication-associated GI and iPART algorithm versus a simple SV count. These results strongly suggest that our approach would work on different cancer types and on other highly remodeled cancer genomes.

### Predictive relevance of iTRAC and iRACIN for chemotherapeutic response in LMS

The overall contribution of curative and adjuvant cytotoxic chemotherapy to 5-year survival in adults was estimated to be 2.3% in Australia and 2.1% in the USA (Morgan, Ward, et Barton 2004). Furthermore, it has been estimated that any class of cancer drugs is ineffective in 75% of patients (Personalized Medicine Coalition. The personalized medicine report Opportunity, Challenges, and the Future 2017; http://www.personalizedmedicinecoalition.org/Userfiles/PMC-Corporate/file/The-Personalized-Medicine-Report1.pdf). Thus, predicting which patients are eligible for which treatment and those who are not is the holy grail of precision medicine. Here we show that chemotherapy for patients with a low iTRAC would be detrimental for their MFS and should therefore be prohibited. In addition, chemotherapy is likely to have no clinical benefit for patients with a medium iTRAC so another therapeutic strategy should be used in them. No conclusion can be drawn for patients with a high iTRAC because of the low statistical power of the log-rank test due to the low number of patients with a high iTRAC which underwent chemotherapy. Nevertheless, high-iTRAC patients would probably benefit from chemotherapy and more inclusions are needed to address this question. On the other hand, iRACIN was not predictively relevant for stratifying the chemotherapeutic response. Nevertheless, we expect it to be relevant for stratifying targeted therapies based on targeting replication and replication-associated repair. We do not consider using MAGIC in this setting as it is a combination of groups of patients albeit similar in their metastatic risk occurrence, but who have different oncogenic mechanisms. Therefore, iPART/iTRAC/iRACIN could serve as a toolset to tackle the question of the relation of GI to therapeutic intervention based on the genomic processes undermining genomic integrity.

### Targeted therapy as a therapeutic strategy in LMS

Our understanding of GI may serve both for prognosis and to orient the choice of therapeutic agent. In patients with germline BRCA1 or 2 mutations, PARP inhibitors have led to major therapeutic advances in patients with ovarian cacner (Moore et al. 2018) and to a lesser extent, breast cancer (Litton et al. 2018) over the past years. Furthermore, it has been proposed that most LMS tumors display hallmarks of “BRCAness”, including alterations in homologous recombination DNA repair genes, enrichment of specific mutational signatures, and cultured LMS cells sensitive to olaparib and cisplatin (Chudasama et al. 2018). Thus, PARP1 inhibitors are a candidate treatment in LMS. The use of iTRAC/iRACIN in clinical settings would allow the selection of LMS patients who potentially would have a significant therapeutic response to PARP1 inhibitors.

### Future directions

Given the far-reaching consequences of GI for treatment success, therapeutic choices and clinical care, an accurate measure of GI and its dynamics is paramount in precision medicine. The development of robust biomarkers enabling GI dynamics to be captured is crucial if we are to leverage the potential of GI for patient stratification purposes and for exploiting this feature for making therapeutic choices. Deriving minimally invasive approaches that enable clinicians to assess whether GI is at play within a given tumor sample might be crucial for its efficient exploitation in clinical settings. Recent advances in whole genome/whole exome sequencing of formalin-fixed, paraffin-embedded (FFPE) tissues allows the application of this approach in routine clinical settings. Finally, prospective findings now need to be validated by measuring iTRAC/iRACIN in circulating tumor DNA (ctDNA) or in circulating tumor cells (CTC) and in subsequent clinical trials.

## MATERIALS and METHODS

### Samples

LMS Samples (112) used in this study were collected as part of the ICGC program (International Cancer Genome Consortium; https://icgc.org/) with patient consent. Samples were frozen tissues provided by pathologists and a blood sample for each included patient provided by medical oncologists. All cases were systematically reviewed by expert pathologists of the French Sarcoma Group according to the World Health Organization recommendations (Fletcher, C. 2013).

### DNA extraction

Genomic DNA from frozen samples was isolated using a standard phenol-chloroform extraction protocol (Chomczynski et Sacchi 1987). DNA was quantified using a Nanodrop 1000 spectrophotometer according to manufacturer’s recommendations (Thermo Scientific, Waltham, MA, USA). Blood material from included patients was also available. Genomic DNA from blood samples was extracted using customized automated purification of DNA from compromised blood samples on the Autopure LS protocol according to the manufacturer’s recommendations (Qiagen, Hilden, Germany), with increased centrifugation of 10 min for DNA precipitation and DNA wash.

### Whole genome sequencing and analysis

To construct short-insert paired-end libraries, a no-PCR protocol was used with the TruSeq™DNA Sample Preparation Kit v2 (Illumina Inc., San Diego, CA, USA) and the KAPA Library Preparation kit (Kapa Biosystems, Basel, Switzerland). Briefly, 2 µg of genomic DNA were sheared on a Covaris™ E220, size-selected and concentrated using AMPure XP beads (Agencourt, Beckman Coulter, Brea, CA, USA) in order to reach a fragment size of 220 – 480 bp. Fragmented DNA was end-repaired, adenylated and ligated to Illumina-specific indexed paired-end adapters.

DNA sequencing was performed in paired-end mode in lanes of HiSeq2000 Flowcell v3 (2×100 bp) or Flowcell v4 (2×125bp) or v4 (2×125 bp) or in sequencing lanes of NovaSeq 6000 Flowcell S4 (2×150bp) (Illumina Inc., San Diego, CA, USA) to analyze tumor or matched normal blood samples (from the same patient) and to reach a minimal yield of 145 or 85 Gb, respectively. Two tumor samples (LMS2T and LMS5T) were sequenced in 20 lanes of HiSeq2000 Flowcell v3 to reach a minimal yield of 560 Gb. Image analysis, base calling and quality scoring of the run were processed using the manufacturer’s software Real Time Analysis (RTA 1.13.48) and followed by generation of FASTQ sequence files by CASAVA (Illumina Inc., San Diego, CA, USA).

DNA reads were trimmed of the 5’ and 3’ low-quality bases (PHRED cut-off 20, maximum trimmed size: 30 nucleotides (nt)) and sequencing adapters were removed with Sickle2 (Joshi NA, Fass JN. (2011). Sickle: A sliding-window, adaptive, quality-based trimming tool for FastQ files (Version 1.33) [Software]. Available at https://github.com/najoshi/sickle)and SeqPrep3 (J. St. John, SeqPrep. (2011) Available at https://github.com/jstjohn/SeqPrep), respectively. Then, DNA curated sequences were aligned using bwa v-0.7.15 (H. Li et Durbin 2009) with default parameters on the Human Genome version hg38 (Schneider et al. 2017) ***(http://genome.ucsc.edu/ ou https://www.ncbi.nlm.nih.gov/grc/human***). Thus, aligned reads were filtered out if their alignment score was less than 20 or if they were duplicated PCR reads, with SAMtools v1.3.1 (Heng Li et al. 2009) and PicardTools v2.18.2 (“Picard Toolkit.” 2019. Broad Institute, GitHub Repository. http://broadinstitute.github.io/picard/; Broad Institute), respectively.

### Random breakage model, Hscore, readable genome size

The readable genome is represented as a single interval of length L in base pairs (bp). The uniform probability Pu of any genomic position to carry a BP is the total number of BP (n) divided by L: Pu=n/L. For a given genomic interval of size (Li) and number of BP (ni), its probability to harbor ni BP under the random breakage model (RBM) is computed by the probability mass function of binomial distribution as: 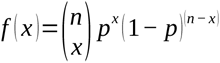; where x=ni, n=Li,p=Pu. The probability of observing more than ni BP under RBM is defined as 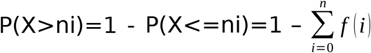; where n = ni, and X is the random variable accounting for the number of observed BP. For the bed file of each DNA element, overlapping intervals were merged (bedtools) and then BP number (ni) and interval size (Li) were computed as follows: ni was computed as the sum of BP in all the merged intervals and Li was computed as the sum of all merged interval sizes. The Hscore is then computed as the -log10 of P(X>ni) under RBM.

We define readable genome size as the total ungapped genome length as defined at https://www.ncbi.nlm.nih.gov/assembly/GCF_000001405.26/. It is equal to 2,948,611,470 bp.

### Breakpoint identification and detection of structural variants

Structural variants (SV) were detected from paired tumor/normal whole genome high-quality sequencing data. Paired-end reads were aligned using Bowtie v2.2.1.0 (Langmead et al. 2009), which is a sensitive local option allowing soft-clipped sequences. The algorithm has three main steps: i) identification of potential breakpoints, ii) characterization of the second side of the breakpoints, and iii) selection of high-confidence breakpoints. All parameters were set to analyze 60X tumor and 30X normal sequencing depth. Very conservative filters were used to minimize false positive detection.

i. Identification: at this step, reads with at least one soft-clipped end were analyzed as singletons. A position was considered as a potential breakpoint if it was covered by at least 4 soft-clipped reads, 5 soft-clipped bases (with at least two occurrences of two different bases), and if they represented more than 5% of the total amount of reads at this position in the tumor sample. We selected potential somatic events by discarding positions covered by at least one read and one base in a surrounding 5-nucleotide window in the normal sample. We refer to them as the “first side” of the breakpoint.
ii. Characterization: to determine the genomic positions of the soft-clipped sequence from selected reads, we used the UCSC blat server (Kent 2002). If no match was returned, the reverse complement sequence was pulled to test. If there was still no match, the BAM file was investigated for some soft-clip somatic position around the discordant or oversized-insert read mate (hereafter named abnormal) location from the first side of the breakpoint. Because of the small size of the soft-clipped sequence, multiple matches can be found. We used soft-clipped abnormal read mates to select matches with the most coherent chromosomic locations. We refer to them as the “second side” of the breakpoint.
iii. Selection: Positions detected from both the first and second sides (in a 5-nucleotide window) were defined as the common pool. We considered as artifacts (due to repeat regions for instance) any couples of positions covered with reads and associated soft-clipped sequences separated by fewer than 15 nucleotides and discarded them. We classified the breakpoints in three groups: high-confidence breakpoints, breakpoints needing investigation, and unique position breakpoints. If a breakpoint was covered by reads and associated soft-clipped sequences having both positions belonging to the common pool, it was classified in the first group. If a breakpoint was covered by reads and associated soft-clipped sequences having only one of the positions belonging to the common pool, it was classified in the second group. Then the missing position was searched among the filtered positions. If it was present in the normal sample, the position was discarded and the breakpoint was completed otherwise. Finally, the third group corresponds to breakpoints with both sides outside the common pool and considered as unique: these were discarded. The sides of breakpoints were sorted according to their chromosomic positions to avoid duplicates.

### Data collection

The following DNA elements were considered in the present analysis: DNA repeats comprising MicroSatellite (MS), Simple Repeats (SR), Low Complexity (LC), Self-Chain segments (SCS) which were classified into self-aligned inverted chains SCS (SCS-S) and gapped SCS (SCS-G), Long Terminal Repeats (LTR), and Retrotransposons (RT); Non-B DNA comprising A-Phased Repeats (APR), Direct Repeats (DR), G-quadruplex (GQ), Inverted Repeats (IR), Mirror Repeats (MR), Short Tandem Repeats (STR), Z-DNA (Z) and R-Loops Forming Sequences (RLFS); and Regulatory DNA elements comprising CpG islands (CpGi), *cis*-regulatory modules (CRM), DNase I hypersensitive site (DHS) of promoter type (DHS_prom), DHS of enhancer type (DHS_enh), DHS of dyadic type (both enhancer and promoter signatures) (DHS_dyadic), and DHS of other types (DHS_rest).

Data for CpG islands, microsatellites, simple repeats, low complexity, retrotransposons, long terminal repeats, self-chains and sequencing gaps were obtained from the UCSC Genome Browser website (http://genome.ucsc.edu/; genome assembly hg38). All Non-B DNA except RLFS were generated using the non-B DNA research tool from the non-B DNA database (Cer et al. 2012). RLFS data were generated using QmRLFS-finder (Jenjaroenpun et al. 2015). CRM data were obtained from Remap2018 (Chèneby et al. 2018) and data were downloaded from (http://pedagogix-tagc.univ-mrs.fr/remap/). DNase I-accessible regulatory regions (with *-log10(p) >= 2)* were downloaded from the roadmap epigenomics project at https://personal.broadinstitute.org/meuleman/reg2map/HoneyBadger2_release/ and coordinates were converted from genome assembly hg19 to hg38 using Liftover, the UCSC coordinates conversion tool (Kuhn, Haussler, et Kent 2013).

### TRAC index/RACIN index

The DNA elements were sorted in separate indexes depending on whether their enrichment in BP was dependent or not on their presence inside or outside the genes (see section below *ingene/outgene split*). DNA elements enriched in BP independently of their position inside or outside the genes were sorted as Replication-Associated Chromosomal INstability elements (RACINe). DNA elements enriched in BP according to their position inside the genes were sorted as TRanscription-Associated Chromosomal instability elements (TRACe). For each TRACe and RACINe element, we pooled all bed files of the corresponding DNA elements into one file and sorted them according to interval positions (bedtools) and merged (bedtools) all overlapping intervals to obtain the corresponding index iTRAC and iRACIN, each as a single bed file. BP counts and interval sizes were computed for each index. These were then used to compute the Hscores under RBM.

### Sliding windows

For each DNA element (sliding window 0 in figures 3, 4 and 5), each genomic feature was shifted (bedtools) by 100 % its length on the positive (+) DNA strand (sliding window +1 in figures 3, 4 and 5) and on the negative (–) DNA strand (sliding window −1 in figures 3, 4 and 5) and the Hscore was computed. This procedure was repeated by shifting each feature ± 2×100 % (sliding window +2, sliding window −2), ± 3x 100 %, until ± 8 100 %.

### Heatmap

The Hscore was computed with Holm’s adjusted p-values procedure (Holm 1979). Two patients (LMS78T and LMS131T) had maximal Hscores of 170.53 and 50.16 while all the other patients had a maximal Hscore less than 39. Therefore, the heatmap was unexploitable as only these two maximal data points were visible. For the sake of clarity, the Hscores of patients LMS78T and LMS131T were normalized by dividing them by 170.53 and multiplying them by 39.

#### Ingene/outgene split

A DNA element was considered as inside a gene if overlapped by at least 1 bp the gene interval delimited by its Transcription Start Site (TSS) and Transcription End Site (TES). Gene coordinates were taken from curated RefSeq entries from the UCSC table browser page (https://genome.ucsc.edu/cgi-bin/hgTables; group=genes and genes prediction;track=NCBI RefSeq; table=RefSeq Curated). Only genes that had expression data in these tumors were considered (for list of genes, see supplemental table 4).

#### SCS-S/SCS-G

Self-chains (SC) were prepared as in (Zhou et al. 2013) except that we split SC segments (SCS) into those are self-aligned (SCS-S) and those are gapped (SCS-G) that is having spacing intervals separting each pair of SC. SCS are defined as the segment of any paired SCs in the same chromosome and their spacing gap. The paired SCs located in different chromosomes and those in the same chromosome but having long spacing intervals (SCS size 30 kb) were filtered out to account only for local interactions. In addition, any SCS-S/SCS-G overlapping with the human genome gaps, SDs was further filtered out.

#### Statistical analysis

All statistical tests and the heatmap were carried out using R (R Core Team (2020);R: A language and environment for statistical computing. R Foundation for Statistical Computing, Vienna, Austria;http://www.r-project.org/index.html).

#### iPART

We consider iPART (Iterative multi-thresholds PARTitioning) to be an unsupervised decision tree (UDT). It is a method that combines the properties and objectives of both unsupervised clustering and decision trees (DT). Hence, iPART looks for thresholds that maximize the differences in groups instead of computing pairwise distances and constructing hierarchical clusters. It resembles DT and regression trees (RT) by using thresholds to split groups. It differs from DT in that it is unsupervised. It also differs from RT in that it does not attempt to predict quantitative variables. The fundamental difference with both RT and DT is its ability to use binary (splitting data into two groups) and ternary (splitting data into three groups) modes, i.e. a crucial feature in our method. It also differs from DT and RT in using the Kaplan-Meier (KM) estimate instead of the GINI purity index or information gain index and sum of squared residuals for DT and RT, respectively. It also resembles unsupervised machine learning like hierarchical clustering and k-means by aiming to find natural patterns/groups in data. On the other hand, it differs from them in that it does not compute pairwise distances or try to construct groups by minimizing their intra-group variance. Instead, it iterates all possible thresholds, find thresholds that maximize the difference between the split groups in terms of the speed at which metastatic events occur in the two groups by minimizing the p-value of the KM test. Briefly, it establishes natural frontiers that maximize the differences between groups instead of constructing groups that minimize intra-group and overall variance.

## Supporting information

LMS cohort characteristics

list of genes used to split DNA elements into ingene vs out genes

Hscore ingenes outgenes and RHscore io

LMS patients clinical data

## Data Availability

all data referred to in the manuscript is available as supplemental tables or described in material and methods section

## Acknowledgment

We are grateful to the French Sarcoma Group for tumor banks and associated clinical annotations and to Jean-Baptiste Courrèges. The following French cancer centers also participated in this study: Centre Paul Papin (Angers), Centre Oscar Lambert (Lille), Institut Paoli Calmettes (Marseille), CHU La Timone (Marseille), CHU Bordeaux and hospital Ambroise Paré (Paris).

This work was supported by grant from the “*Instituts Thematiques Multiorganismes*” (ITMO) Cancer (INSERM and INCa) and the Claudius Regaud Institute.

The authors would like to thank the Centre Nacional d’Anàlisi Genòmica (CNAG, Barcelona, Spain) for WG sequencing services. We thank GENOTOUL for bioinformatics facilities and computer farm.

