## Supplementary material for "Metastatic risk stratification of leiomyosarcoma patients using transcription- and replication-associated chromosomal instability mechanisms": LMS cohort characteristics

| **Characteristics** | **LMS cohort (n=112)** |
| --- | --- |
| **Median follow-up (months)[s.d]** | 47.24[79.70] |
| **Median age (years)[s.d]** | 64[12.01] |
| **Female sex ( %)** | 88(78) |
| **FNCLCC grade ( %)** |  |
| 1 and 2 | 56(50) |
| 3 | 48(43) |
| ND | 8(7) |
| **CINSARC ( %)** |  |
| C1 | 28(25) |
| C2 | 76(68) |
| ND | 8(7) |
| **Histological type ( %)** |  |
| Leimyosarcomas | 70(62) |
| Differentiated | 32(29) |
| Poorly differentiated | 10(9) |
| **Location ( %)** |  |
| Gyneacological area | 15(13) |
| Trunk wall | 7(6) |
| Extremetes | 23(21) |
| Internal trunk | 58(52) |
| others | 9(8) |
| **Median size (mm)** | 80(72) |
| **Depth of tumor ( %)** |  |
| Deep | 93(83) |
| Superficial | 11(10) |
| Superficial and deep | 8(7) |
| **Metastatic events(%)** |  |
| Yes | 53(47) |
| No | 59(53) |
| **Therapeutic management(%)** |  |
| Surgery | 109(97) |
| Surgery + radiotherapy | 35(31) |
| Surgery + chemotherapy | 18(16) |
| Surgery + radiotherapy + chemotherapy | 9(8) |
| Missing data | 3(3) |

**Supplemental table 1 : LMS patients characteristics**

ND : Not determined

Sd : standard deviation
